# Low SARS-CoV-2 sero-prevalence based on anonymized residual sero-survey before and after first wave measures in British Columbia, Canada, March-May 2020

**DOI:** 10.1101/2020.07.13.20153148

**Authors:** Danuta M Skowronski, Inna Sekirov, Suzana Sabaiduc, Macy Zou, Muhammad Morshed, David Lawrence, Kate Smolina, May A Ahmed, Eleni Galanis, Mieke N Fraser, Mayank Singal, Monika Naus, David M Patrick, Samantha E Kaweski, Christopher Mill, Romina C Reyes, Michael T Kelly, Paul N Levett, Martin Petric, Bonnie Henry, Mel Krajden

## Abstract

**Background:** The province of British Columbia (BC) has been recognized for successful SARS-CoV-2 control, with surveillance data showing amongst the lowest case and death rates in Canada. We estimate sero-prevalence for two periods flanking the start (March) and end (May) of first-wave mitigation measures in BC.

**Methods:** Serial cross-sectional sampling was conducted using anonymized residual sera obtained from an outpatient laboratory network, including children and adults in the Greater Vancouver Area (population ∼3 million) where community attack rates were expected to be highest. Screening used two chemiluminescent immuno-assays for spike (S1) and nucleocapsid antibodies. Samples sero-positive on either screening assay were assessed by a third assay targeting the S1 receptor binding domain plus a neutralization assay. Age-standardized sero-prevalence estimates were based on dual-assay positivity. The May sero-prevalence estimate was extrapolated to the source population to assess surveillance under-ascertainment, quantified as the ratio of estimated infections versus reported cases.

**Results:** Serum collection dates spanned March 5-13 and May 15-27, 2020. In March, two of 869 specimens were dual-assay positive, with age-standardized sero-prevalence of 0.28% (95%CI=0.03-0.95). Neither specimen had detectable neutralizing antibodies. In May, four of 885 specimens were dual-assay positive, with age-standardized sero-prevalence of 0.55% (95%CI=0.15-1.37%). All four specimens had detectable neutralizing antibodies. We estimate ∼8 times more infections than reported cases.

**Conclusions:** Less than 1% of British Columbians had been infected with SARS-CoV-2 when first-wave mitigation measures were relaxed in May 2020. Our findings indicate successful suppression of community transmission in BC, but also substantial residual susceptibility. Further sero-survey snapshots are planned as the pandemic unfolds.

**Key points:** Cross-sectional sampling of anonymized residual sera at the start and end of first-wave mitigation measures in British Columbia, Canada shows SARS-CoV-2 sero-prevalence below 1% throughout the winter-spring 2020. Findings indicate successful suppression of community transmission but also substantial residual susceptibility.

## Introduction

British Columbia (BC) is the third-most populous and westernmost province of Canada, with more direct flights from mainland China to the Vancouver International Airport than any other airport in North America or Europe. In that context, BC health officials have long prepared for the potential introduction of emerging respiratory pathogens, with prior success in the early detection and containment of imported human cases of severe acute respiratory syndrome coronavirus (SARS-CoV-1) in 2003 and avian influenza H7N9 in 2015 [1,2].

When on December 31, 2019 a cluster of SARS-like illness became known in Wuhan City, China, the BC Centre for Disease Control (BCCDC) responded rapidly, disseminating provincial alerts that morning and repeatedly over the ensuing weeks. Ultimately, the Wuhan cluster became the coronavirus infectious disease 2019 (COVID-19) pandemic attributed to novel SARS-CoV-2. Although no airport in Canada receives direct flights from Wuhan, the earliest COVID-19 onset date in Canada (January 15) was a case arriving indirectly from Wuhan to Vancouver, with the earliest reported cases otherwise in Canada also among travelers arriving indirectly from Wuhan to Toronto, Ontario (January 25) or Vancouver, BC (January 27). Thereafter, BC also reported the first importation of COVID-19 from outside China (Iran) on February 20; the first super-spreading event on March 6; and the first long-term care facility outbreak (and associated death) on March 9 [3].

Despite these firsts, BC has been recognized for its successful control of COVID-19 during the winter-spring 2020. Early success was attributed in part to timely alerting; sustained media messaging through a single health official; rapid development and deployment of diagnostic testing; and restriction of staff movement between long-term care facilities. As elsewhere in Canada, on March 12 the province recommended against non-essential travel that preceded the March 16 start of the usual two-week spring-break for schools; whereas, in other provinces (e.g. Quebec), spring break began two weeks earlier (March 2) and families were already returning from trips abroad. In BC, a period of population-level mitigation measures, incorporating further travel restrictions, bans on public gatherings and closure of personal service establishments ultimately spanned ∼2 months, with re-opening beginning gradually by May 19. This period captured the beginning and end of peak COVID-19 case reporting, corresponding with first wave activity in BC (**Figure 1**) [3,4].

**Figure 1.**
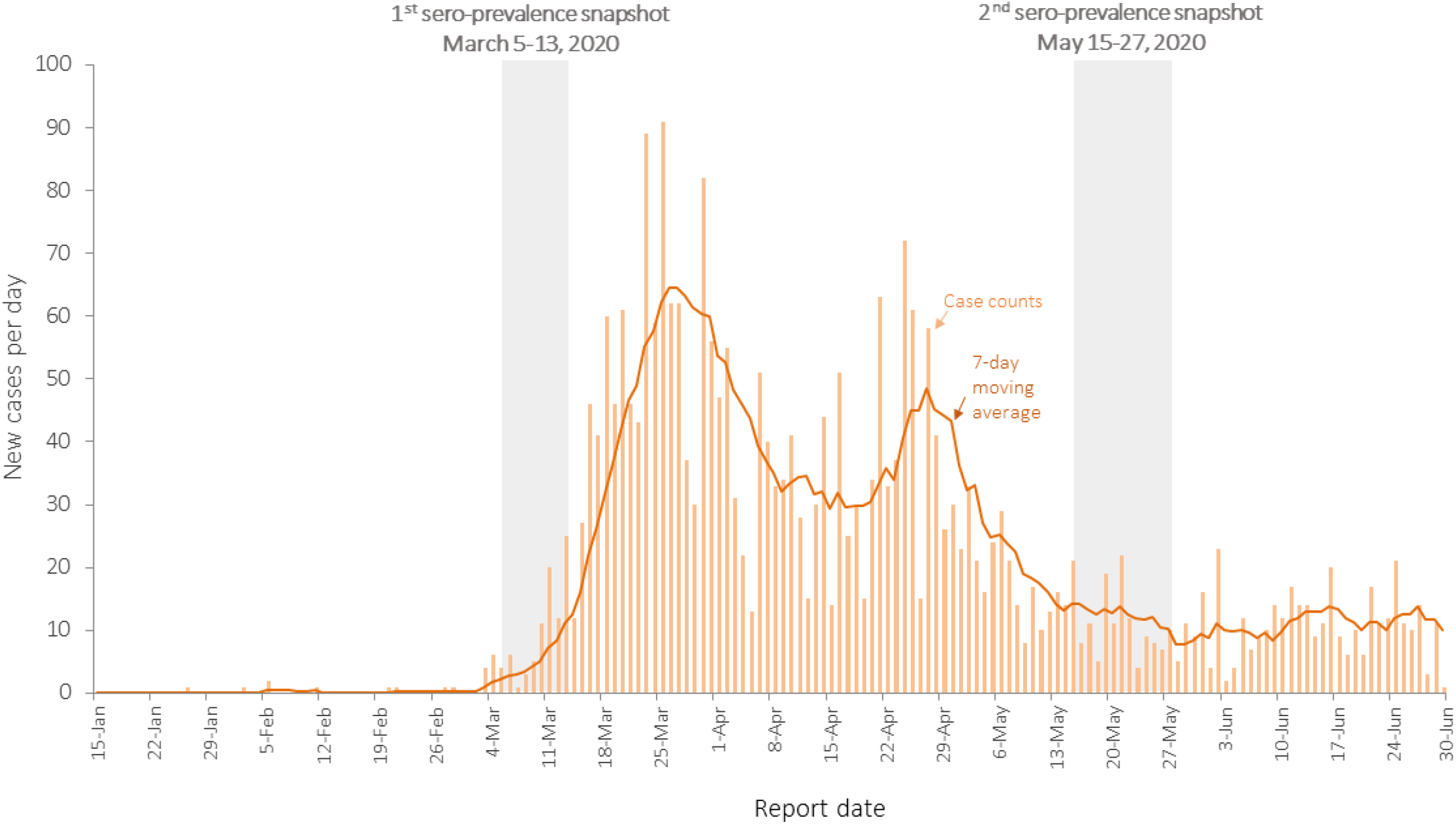
Provincial epidemic curve of incident cases by report date, British Columbia (BC), January 15 to June 30, 2020 (N=2,916) Displayed are the daily number of new cases by report date for the province of BC, plotted as case counts (bars) and 7-day moving average (line). Super-imposed is the span of March and May serum collection dates.

Although other areas enacted similar control measures, BC has consistently reported amongst the lowest COVID-19 case and death rates compared to other provinces in Canada, adjacent west coast states in the United States (US) (**Figure 2)** [5,6], or elsewhere globally [4,7]. Until early April, however, BC targeted SARS-CoV-2 testing to people with exposure and/or high-risk indication, leading to a per capita testing rate that was also below other provinces [4,7]. Like elsewhere, all-cause deaths in BC exceeded tallies of the previous 5 years (during a six-week period spanning March 15 to April 25) [8,9]. Taken together these statistics have led to speculation that BC may have experienced more undocumented SARS-CoV-2 infections that, in conjunction with asymptomatic cases, contributed to unrecognized community transmission [10,11].

**Figure 2.**
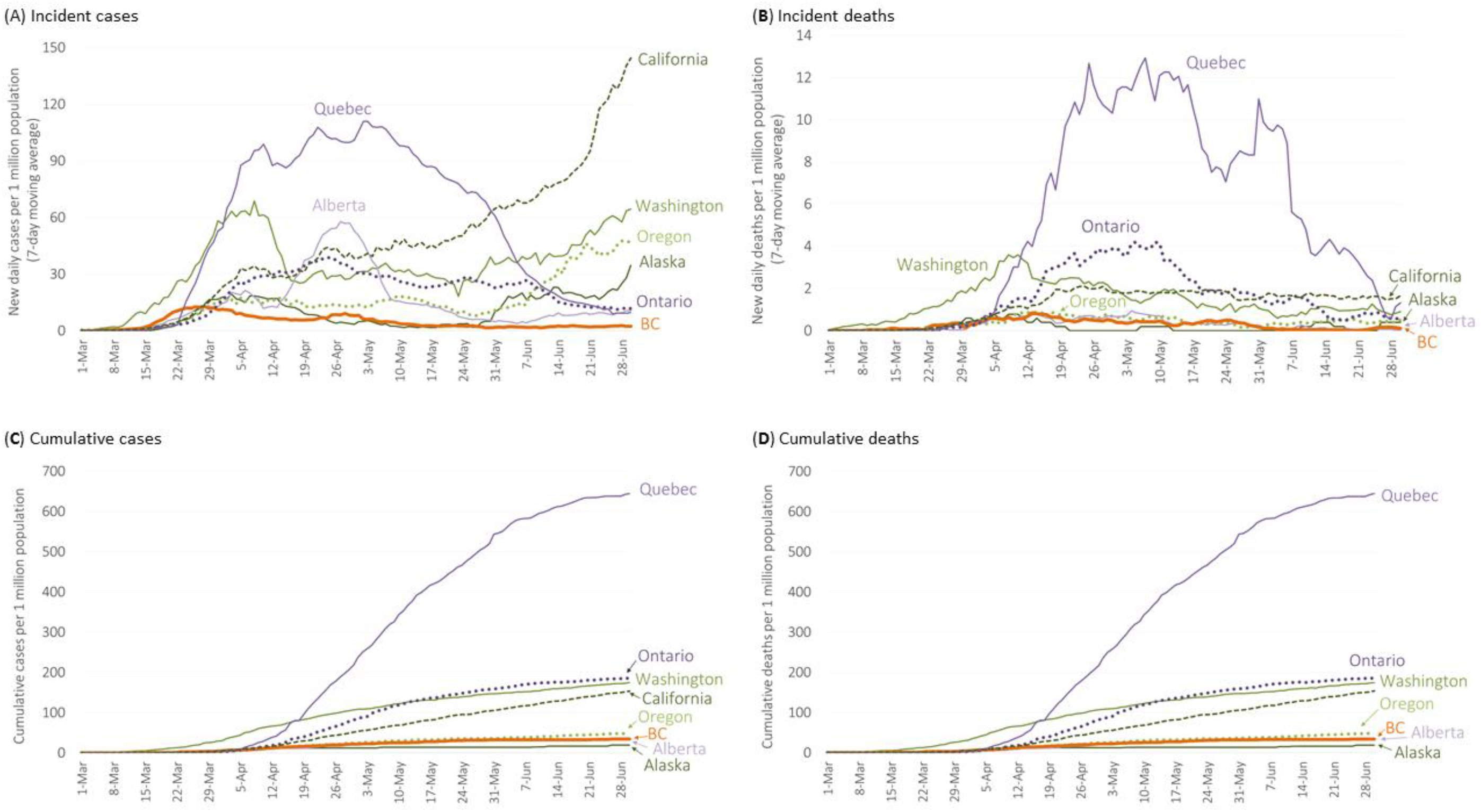
Daily incident and cumulative COVID-19 case and death rates, British Columbia (BC) and comparator jurisdictions, March 1 to June 30, 2020. Displayed are daily COVID-19 rates per million population shown by report date for (A) incident cases; (B) incident deaths; (C) cumulative cases; and (D) cumulative deaths comparing BC to the three other most populous provinces of Canada (Alberta, Ontario and Quebec) and other west coast United States (Alaska, California, Oregon, Washington states). All 8 jurisdictions reached a cumulative incidence of 2 per million between March 2 and 17, 2020. COVID-19 cases for BC, Alberta, Ontario and Quebec reflect daily public briefings by respective governments. COVID-19 confirmed cases for Alaska, California, Oregon and Washington come from the COVID-19 Data Repository of the Center for Systems Science and Engineering at Johns Hopkins University (https://github.com/CSSEGISandData/COVID-19). Population data for Canadian provinces are from Statistics Canada [5] and for the US are from the United States Census Bureau [6].

As part of risk assessment for other emerging respiratory pathogens, the BCCDC has previously established a sero-survey protocol that utilizes anonymized residual patient specimens. Sampling is strategically targeted to the Lower Mainland, BC (i.e. Greater Vancouver Area, including the Fraser Valley) [2,12-17], where ∼60% of the provincial population resides [18], and where community attack rates are expected to be highest. Using this protocol, the BCCDC has previously monitored attack rates, cumulative incidence, and residual susceptibility by age group across successive waves of the influenza A(H1N1)pdm09 pandemic [12-15], and has contributed findings to meta-analyses of the World Health Organization (WHO) [15].

In February 2020, BCCDC adapted this protocol for SARS-CoV-2 assessment. We present baseline findings for two cross-sectional serum collections (or snapshots) flanking the start (March) and end (May) of first wave activity and population-level control measures. The goal of these serial snapshots was to establish baseline and early pandemic sero-prevalence for future attack rate comparison; to estimate cumulative incidence, residual susceptibility and the extent to which community transmission was suppressed; and to assess surveillance under-ascertainment across the winter-spring 2020 period in BC.

## Methods

### Sampling

As shown in **Figure 1**, the first snapshot captured specimens collected March 5-13 and the second snapshot captured specimens collected May 15-27, 2020. For each snapshot, at least 500μL of anonymized residual sera were obtained from patients attending one of ∼80 diagnostic service centres of the only outpatient laboratory network in the Lower Mainland, BC (LifeLabs). Specimens were collected as shown geographically distributed across this region in **Figure 3** [18]. Consistent with the WHO Unity protocol [19], sera were obtained from 100 patients within each of the following age groups (equally male and female): <5, 5-9, 10-19, 20-29, 30-39, 40-49, 50-59, 60-69, 70-79, ≥ 80 years. Accompanying detail included age in years, sex and city. Specimens were included only if at least 200μL of residual sera were to remain after testing. The sero-survey was authorized by the Provincial Health Officer and approved by the Clinical Research Ethics Board of the University of British Columbia (H20-00653).

**Figure 3.**
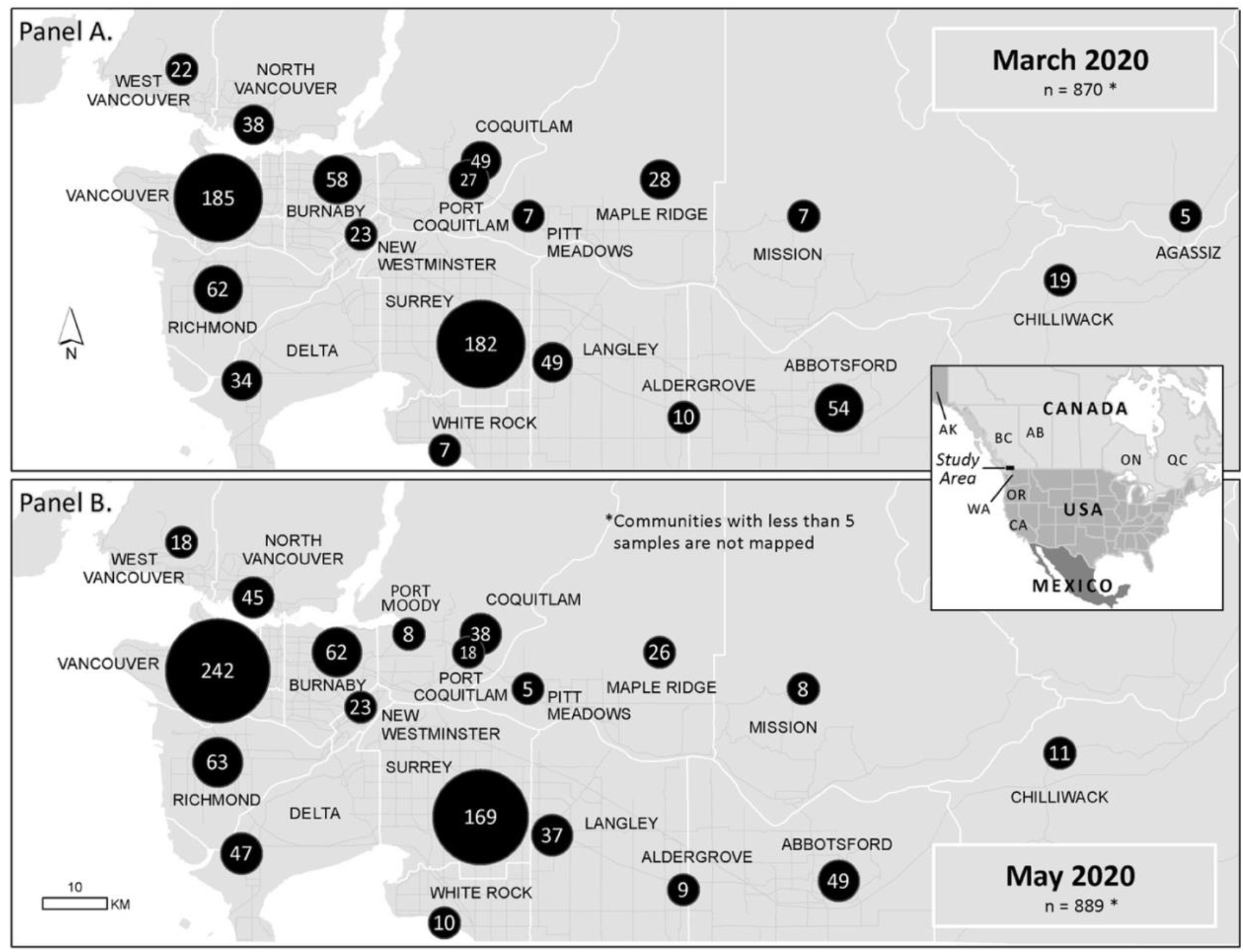
Distribution of included serum samples by community of collection across the Lower Mainland, British Columbia (BC), March and May, 2020 snapshots. Panels display the number of serum samples by community of collection included in serological testing by either screening assay for the March (Panel A) and May (Panel B) snapshots. Communities contributing less than 5 samples are not mapped. The March snapshot included 870 samples tested for spike (S1) and 869 tested for nucleocapsid antibodies; the May snapshot included 889 samples tested for S1 and 885 tested for nucleocapsid antibodies. Shown in Canada (west to east): BC=British Columbia, Canada; AB=Alberta; ON=Ontario; QC=Quebec. Shown in the United States (north to south): AK=Alaska; WA=Washington State; OR=Oregon; CA=California.

### Serological testing

Sera were screened by two chemiluminescent immuno-assays (CLIA). Specimens sero-positive on either screening assay were subjected to a third CLIA as well as neutralization assay.

Screening assays, both currently approved by Health Canada [20], included:

1. Detection of total antibody (IgA, IgG and IgM) to recombinant spike (S1) protein using the Vitros XT 7600 analyzer (Ortho-Clinical Diagnostics, Rochester, New York) [21]. The patient sample signal was divided by the calibrator signal, with resultant signal to cut-off (S/C) ratios of <1.00 and ≥1.00 considered negative or positive, respectively. Public Health England reports overall sensitivity of 85% (95% confidence interval (CI)=76.5-91.4) for this assay, 91.8% (95%CI=83.8-96.6) at ≥14 days and 93.5% (95%CI=85.5-97.9) at ≥21 days after symptom onset, with specificity of 99.5% (95%CI=98.2-99.9) [22].
2. Detection of IgG antibody to nucleocapsid using the ARCHITECT i2000SR analyzer (Abbott Laboratories, Diagnostic Division, Abbott Park, Illinois) [21]. Samples with S/C ratios of <1.40 and ≥1.40 were considered negative or positive, respectively. Public Health England reports overall sensitivity of 92.7% (95%CI=85.6-97.0) for this assay, 93.9% (95%CI=86.3-98.0) at ≥14 days and 93.5% (95%CI=85.5-97.9) at ≥21 days after symptom onset, with specificity of 100% (95%CI: 99.1-100) [22]. Theel et al. report sensitivity of 97.3% (95%CI=85-100) at ≥15 days with specificity of 99.6% (95%CI: 97.6—100) [23]. Bryan et al. report sensitivity of 82.4% (95%CI=51.0-76.4) at ≥10 days, 96.9% (89.5-99.5) at ≥14 days and 100% (95.1-100) at ≥17 days after symptom onset [24]. Further testing of sero-positive specimens included:
3. Detection of total (IgG, IgM) antibodies to the S1 receptor-binding domain (S1-RBD) by the ADVIA Centaur XPT system (Siemens Healthineers, Erlangen, Germany) [21]. Samples with S/C ratios of <1.0 and ≥1.0 were considered negative or positive, respectively. As of July 1, 2020, this assay had not received Health Canada approval but has FDA Emergency Use Authorization [20,21]. For a similar Siemens assay based upon the Atellica-IM analyzer, Public Health England reports overall sensitivity of 86.0% (95%CI=77.6-92.1), 89.4% (95%CI=80.8-95.0) at ≥14 days and 92.4% (95%CI=84.2-97.2) at ≥21 days after symptom onset, with specificity of 100% (95%CI=99.1-100) [22].
4. Standard virus neutralization assay as described by Zakhartchouk et al. [25]. Each serum specimen was heat inactivated at 56°C for 30min and duplicate serial 2-fold dilutions from 1:8 to 1:4096 were each incubated with 100 TCID^50^ of SARS-CoV-2 for 2h, then added to monolayers of Vero-E6 cells. Seed virus was obtained from the National Microbiology Laboratory designated as SARS-CoV-2/2020/0044. Monolayers were examined after 72h for characteristic cytopathogenic effect (CPE). The inverse of the highest serum dilution to inhibit CPE was deemed the antibody titre. Geometric mean titres (GMT) of duplicate tests are presented.

### Sero-prevalence assessment

The number and proportion of specimens testing positive by each screening assay are presented by age group and snapshot with 95%CIs derived by exact method. We explored the proportion sero-positive on either screening assay and further taking into account sensitivity of 85% independently applied to each screening assay while assuming perfect specificity (increasing the tally sero-positive on either screening assay by a factor of 1/0.85). Primary sero-prevalence estimates are defined by sero-positivity on at least two of the four testing assays. Sero-prevalence estimates were age-standardized (direct method) to the 2020 Lower Mainland source population [18], with 95%CI derived using the method proposed by Tiwari et al. based upon gamma approximation [26].

To assess the order of magnitude of surveillance under-ascertainment, we derived a ratio of estimated infections to reported cases. The estimated number of infections was calculated as the product of the age-standardized point estimate of sero-prevalence from the May snapshot multiplied by the approximate source population size (3 million). Taking into account variable delay from infection to detectable antibody response (∼1-2 weeks) [27] and from case detection to surveillance reporting, we also approximated the number of reported cases. Cumulative tallies were obtained for combined Vancouver Coastal and Fraser Health Authorities from on-line surveillance reports for May 1 (n=1801), May 8 (n=1955), May 15 (n=2042) and May 27 (n=2166) [4], summarized as ∼2000 reported cases for this analysis.

## Results

### Overall profile

There were 1759 sera included in both snapshots; 241 specimens were set aside because of low sample volume of which 163 (68%) were children 0-4 years and 39 (16%) were children 5-9 years. For the March snapshot, 870 and 869 sera were screened for antibodies to S1 and nucleocapsid, respectively; for the May snapshot 889 and 885, respectively, were tested (**Table 1**). At each snapshot, median age (45 and 45 years, respectively) and the proportion female (51% and 50%, respectively) were comparable to the Lower Mainland source population (40 years and 51%, respectively) [18].

**Table 1.** Results of SARS-CoV-2 sero-survey screening by chemiluminescent assay for antibodies to spike (S1) and nucleocapsid proteins, by age group, March and May 2020 snapshots, Lower Mainland, BC, Canada

| Age group (years) | March 2020 Snapshot |  |  |  |  | May 2020 Snapshot |  |  |  |  |
| --- | --- | --- | --- | --- | --- | --- | --- | --- | --- | --- |
|  | N | Median age (years) | Female Number (%) | Spike S1 Positive N (%; 95% CI) | Nucleocapsid Positive N (%; 95% CI) | N | Median age (years) | Female Number (%) | Spike S1 Positive N (%; 95% CI) | Nucleocapsid Positive N (%; 95% CI) |
| <10 | 96<sup>a</sup> | 6 | 52 (54) | 0 | 0/95<sup>b</sup> | 102<sup>c</sup> | 7 | 52 (51) | 0 | 0 |
| 10-19 | 83 | 16 | 47 (57) | 0 | 0 | 94 | 17 | 46 (49) | 1<br>(1.1; 0.03 – 5.8) | 1/93<sup>b</sup><br>(1.1; 0.03 – 5.9) |
| 20-29 | 98 | 25 | 50 (51) | 0 | 1<br>(1.0; 0.03 – 5.6) | 98 | 25.5 | 50 (51) | 0 | 0 |
| 30-39 | 97 | 35 | 48 (49) | 0 | 0 | 100 | 34 | 50 (50) | 1<br>(1.0; 0.03 – 5.5) | 1<br>(1.0; 0.03 – 5.5) |
| 40-49 | 100 | 44 | 50 (50) | 1<br>(1.0; 0.03 – 5.5) | 0 | 99 | 45 | 50 (51) | 0 | 0 |
| 50-59 | 99 | 55 | 50 (51) | 2<br>(2.0; 0.25 – 7.1) | 0 | 98 | 54 | 49 (50) | 3<br>(3.1; 0.64 – 8.7) | 3<br>(3.1; 0.64 – 8.7) |
| 60-69 | 100 | 64 | 50 (50) | 1<br>(1.0; 0.03 – 5.5) | 0 | 100 | 64.5 | 50 (50) | 0 | 0 |
| 70-79 | 100 | 73 | 50 (50) | 1<br>(1.0; 0.03 – 5.5) | 1<br>(1.0; 0.03 – 5.5) | 100 | 74 | 50 (50) | 1<br>(1.0; 0.03 – 5.5) | 1/99<sup>b</sup><br>(1.0; 0.03 – 5.5) |
| 80+ | 97 | 85 | 47 (48) | 2<br>(2.1; 0.25 – 7.3) | 2<br>(2.1; 0.25 – 7.3) | 98 | 85.5 | 50 (51) | 0 | 1/96<sup>d</sup><br>(1.0; 0.03 – 5.7) |
| **TOTAL (crude)** | 870 | 45 | 444 (51) | 7/870<br>(0.80; 0.32 – 1.65) | 4/869<sup>b</sup><br>(0.46; 0.13 – 1.17) | 889 | 45 | 447 (50) | 6/889<br>(0.67; 0.25 – 1.46) | 7/885<sup>e</sup><br>(0.79; 0.32 – 1.62) |

<sup>a</sup> Of 200 sera collected from children 0-4 and 5-9 years of age, sufficient sera were available for this initial serological analysis for 25/101 and 71/99, respectively. Most included children < 10 years old were therefore 5-9 years of age (i.e. 71/96; 74%).
<sup>b</sup> One specimen was tested for antibodies to S1 but not nucleocapsid because below the minimum sample volume established by our protocol
<sup>c</sup> Of 200 sera collected from children 0-4 and 5-9 years of age, sufficient sera were available for this initial serological analysis for 13/100 and 89/100, respectively. Most children < 10 years old were therefore 5-9 years of age (i.e. 89/102; 87%).
<sup>d</sup> Two specimens were assessed for antibodies to S1 but not nucleocapsid because below the minimum sample volume established by our protocol
<sup>e</sup> Four specimens in total from the May snapshot were assessed for antibodies to S1 but not nucleocapsid because below the minimum sample volume established by our protocol

### March snapshot

Among specimens included from the March snapshot, 7/870 (0.80%; 95%CI=0.32-1.65) were S1 sero-positive and an additional 4/869 (0.46%; 95%CI=0.13-1.17) were nucleocapsid positive (**Table 1**). Eleven of 869 (1.27%; 95%CI=0.63-2.25) specimens were positive on either assay (none from children <20 years old) (**Table 2**), with age-standardized proportion of 1.01% (95%CI=0.46-1.92). Accounting for imperfect test sensitivity of 85%, the number positive on either screening assay might have been 13 of 869 or up to 1.50% (95%CI=0.80-2.54). However, no March specimens were positive on both screening assays and only when additionally assessed on the third CLIA, did 2 specimens show dual-assay positivity, to S1 and S1-RBD. Both were identified among adults 40-59 years of age (2/199; 1.01%; 95%CI=0.12-3.58). None of the sero-positive specimens in March had detectable neutralizing antibodies (**Table 2**).

**Table 2.**
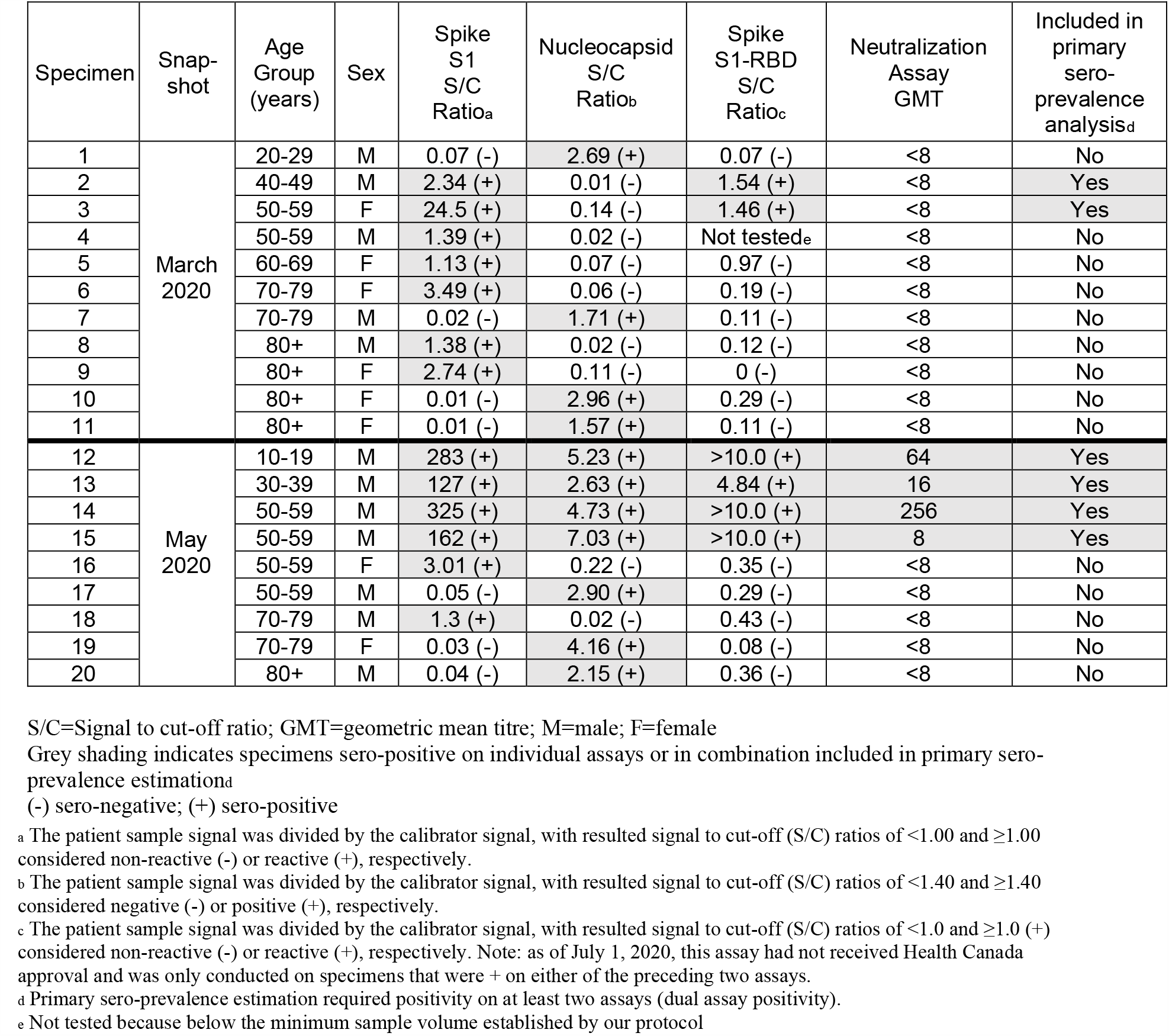
Specimens sero-positive on either spike S1 or nucleocapsid screening assay, including additional S1 receptor binding domain and neutralization assay findings, March and May 2020

Two of 869 sera were dual-assay positive at the March snapshot giving a crude sero-prevalence of 0.23% (95%CI=0.03-0.83) and age-standardized sero-prevalence of 0.28% (95%CI=0.03-0.95).

### May snapshot

Among specimens included from the May snapshot, 6/889 (0.67%; 95%CI=0.25-1.46) were S1 sero-positive and 7/885 (0.79%; 95%CI=0.32-1.62) were nucleocapsid positive (**Table 1**). Nine of 885 (1.02%; 95%CI=0.47-1.92%) specimens were positive on either assay (**Table 2**), with age-standardized proportion of 1.03% (95%CI=0.45-2.00). Accounting for imperfect test sensitivity of 85%, the number positive on either screening assay might have been 11 of 885 or up to 1.24% (95%CI=0.62-2.21).

Four specimens were positive on both S1 and nucleocapsid screening assays and when additionally tested the same four specimens were also S1-RBD and neutralization assay positive; whereas, the 5 specimens positive on just one or the other screening assay were neither S1-RBD nor neutralization assay positive (**Table 2**). The four consistently positive sera included 1/195 children <20-years-old (0.51%; 95%CI=0.01-2.82) and 3/297 adults 30-59-years-old (1.01%; 95%CI=0.21-2.92). The S1, nucleocapsid and S1-RBD signals on these four specimens were each high at ≥127, ≥2.63 and ≥4.84, respectively. The two sera with the highest S1 signals (283 and 325, respectively) and S1-RBD signals (>10.0) also had the highest neutralizing antibody titres (GMTs of 64 and 256, respectively) although one patient with strong S1, nucleocapsid, and S1-RBD signals (162, 7.03, >10.0) had only low-level neutralization titre (GMT of 8).

Four of 885 sera were dual-assay positive at the May snapshot giving a crude sero-prevalence of 0.45% (95%CI=0.12-1.15) and age-standardized sero-prevalence of 0.55% (95%CI=0.15-1.37%). Applying this 0.55% age-standardized sero-prevalence point estimate to the Lower Mainland source population (∼3 million), we estimate about 16,500 infections, or about 8 times higher than the ∼2000 cases reported.

## Discussion

We report the first SARS-CoV-2 sero-prevalence estimates from Canada. Our estimates are based on anonymized residual sera obtained from an outpatient laboratory network in an area of British Columbia where community attack rates were anticipated to be highest. Serial sampling captured two periods flanking the start and end of the first pandemic wave and the implementation of population measures to mitigate its impact. Using a convenient and efficient sero-survey protocol previously used by the BCCDC for emerging pathogen risk assessment [12-17], we estimate SARS-CoV-2 cumulative incidence (community-wide infection rates) remained below 1% throughout the winter-spring 2020 period in BC.

For efficiency in serological testing we used commercially-available, high-throughput CLIAs and for improved reliability applied an algorithm involving multiple viral targets. This included two screening assays, one targeting the S1 and another the nucleocapsid, with a third CLIA applied to positive specimens that targeted the S1-RBD [20,21]. All positive specimens were further assessed by gold-standard neutralization assay [25]. We estimated sero-prevalence based on dual-assay positivity and report cumulative incidence of 0.28% by the start of first wave population-level measures in March. Had we interpreted sero-positivity based on either screening assay, the age-standardized proportion would have just reached 1.0% and taking into account test sensitivity as low as 85%, sero-positivity might have reached 1.5%. In fact, just two of 869 March specimens were sero-positive to both S1 and S1-RBD, but none were sero-positive on both S1 and nucleocapsid screening assays and none had detectable neutralizing antibodies. In that regard, the only two dually-positive specimens in March may have been false-positives and community-level attack rates could be even lower than we estimate by that snapshot. Potential cross-reactivity between SARS-CoV-2 and other coronaviruses may explain inconsistent findings in March and will be explored in follow-up investigations [27,28]. Alternatively, SARS-CoV-2 cases, notably mild or asymptomatic infections, may not have mounted a serological response or sustained neutralizing titres [29-31]. A similar inability to detect neutralizing antibodies among specimens positive on enzyme immunoassay was also recently reported from Hong Kong [32].

By the end of the first pandemic wave and easing of restrictions in May, we estimate sero-prevalence and cumulative incidence remained low at 0.55%. Had we interpreted sero-positivity based on either screening assay, the age-standardized proportion would have still been just 1.0%, and accounting for a test sensitivity of 85%, sero-positivity might have reached 1.2%. Unlike the earlier March snapshot, several May specimens showed S1 plus nucleocapsid positivity, and the same four specimens were also S1-RBD and neutralization assay positive, confirming true infections. Overall, just 20 of 1754 screened sera overall were positive on any assay (1.14%;95%CI=0.70-1.76), a low proportion indicating that if non-specific reactivity or cross-reactivity with other endemic coronaviruses is an issue [27,28], it is not a prominent one. Furthermore, all sera that were positive for S1-RBD were also positive for S1 antibodies; and all sera that were positive for S1 and that had a neutralizing antibody response were also nucleocapsid sero-positive. In combination, these findings highlight that in lieu of labour-intensive screening by gold-standard neutralization assay, an efficient algorithm for sero-prevalence estimation might rely upon orthogonal (two-tiered) high-throughput CLIA testing [33]. In particular, assays targeting S1 or S1-RBD, plus nucleocapsid, should provide reliable and specific detection of SARS-CoV-2 sero-positivity. Such orthogonal approach, however, requires further validation.

Multiple other countries have reported sero-prevalence estimates [24,32,34-40], but study populations, sampling approach, time frame, as well as antibody assays and algorithms must be taken into account when comparing findings. Survey approaches based on blood donation, invitation or other voluntary participation may over-estimate sero-prevalence due to self-selection bias. Few other studies have capitalized on residual patient specimens to try to mitigate that bias [32,37,39], and in that regard, our findings are best compared to another such study recently reported from the US involving children and adults across six cities [39]. For the period March 23-April 1, US authors reported age-standardized sero-prevalence ranging from 1.13% (95%CI=0.70-1.94) for the Puget Sound Region of Washington state to 6.93% (95%CI=5.02-8.92) for New York City [39]. Note that the latter estimate using residual patient specimens is substantially lower than estimated by invitation to adult grocery store patrons in New York City between April 19-28 (22.7%;95%CI=21.5-24.0) [40]. For the Puget Sound Region, US authors estimated about 11 times as many infections compared to reported cases [39], a comparable order of magnitude to our estimate of about 8-times as many infections.

There are limitations to this analysis. Delay in generating an antibody response and uncertainty in the duration of antibody persistence, particularly among mild or asymptomatic cases and for neutralizing titres [27,29-31], may lead to under-estimation of sero-prevalence. Specimens were anonymized without accompanying clinical detail such as symptom history or testing indication. We restricted to outpatient specimens and show both snapshots to be generally representative of the source population for age and sex. However, residual clinical specimens are more likely to come from people with underlying comorbidity and may differ from the rest of the population in their exposure risk, immune response and healthcare seeking. We assume sampling within the Lower Mainland provides an upper range of community sero-prevalence for BC; however, this does not preclude discrete pockets experiencing higher attack rates. Given manifold uncertainties, our ratio of estimated infections versus reported cases is an imprecise approximation, meant only to gauge the likely order of magnitude of surveillance under-ascertainment. In addition to other considerations, reported surveillance tallies may include cases that were imported or accrued within care facilities or other settings under-represented in our community-based sero-sampling. Finally, with such low sero-prevalence, including just one infected child in 375 pediatric sera tested, we were limited in our ability to conduct subset comparisons.

In conclusion, we present a convenient, efficient and reliable approach for serial population-based sero-prevalence monitoring based on anonymized residual sampling (mitigating self-selection bias) and orthogonal high-throughput testing (improving predictive value). We estimate <1% of British Columbians were infected with SARS-CoV-2 by the time first wave restrictions were relaxed in May. These sero-prevalence findings reinforce other surveillance data indicating successful suppression of SARS-CoV-2 transmission throughout the winter-spring 2020 period in BC. This success, however, constitutes a double-edged sword, further highlighting substantial residual susceptibility. Our sero-prevalence protocol is readily amenable to comparison across serial snapshots and these are planned at relevant intervals as the pandemic unfolds.

## Data Availability

Data requests may be directed to the corresponding author and will be made available in accordance with the conditions of research ethics board  and provincial privacy review.

## Acknowledgments

Authors wish to acknowledge the serological testing contribution of the following individuals at the British Columbia Centre for Disease Control (BCCDC) Public Health Laboratory (PHL): Tamara Pidduck, Annie Mak, Jesse Kustra, Chris Ikeda, Joanne Cho, Damien Vilaysane, Joseph Cuison, Kyra Laureano, Mo Ng, Navdeep Chahil, Jonathan Laley, Erin Caruthers, Lori Willis, Irene Yan, Fred Burgess, Ray Wada, Leanne Gagnier, Rosemarie Blancaflor, Christine Froehlich, and Sara Chung. We thank Rhonda Creswell of LifeLabs for diligent supervision of serum collection and Shinhye Kim of the BCCDC for coordination of this research. Finally, we thank the many frontline, regional and provincial practitioners, including clinical and public health providers, epidemiologists, Medical Health Officers and others for their significant contributions to surveillance case reporting and COVID-19 control measures in British Columbia.

## References

1. Skowronski DM, Petric M, Daly P, et al. Coordinated response to SARS, Vancouver, Canada. Emerg Infect Dis, 2006;12:155–8.

2. Skowronski DM, Chambers C, Gustafson R, et al. Avian Influenza A(H7N9) Virus Infection in 2 Travelers Returning from China to Canada, January 2015. Emerg Infect Dis, 2016;22:71–4.

3. Government of British Columbia. COVID-19 (Novel Coronavirus). Available at: https://www2.gov.bc.ca/gov/content/health/about-bc-s-health-care-system/office-of-the-provincial-health-officer/current-health-topics/covid-19-novel-coronavirus Accessed 8 July 2020.

4. British Columbia Centre for Disease Control. COVID-19 Surveillance Reports. Available at: http://www.bccdc.ca/health-info/diseases-conditions/covid-19/data. Accessed 8 July 2020.

5. Statistics Canada. Table 17-10-0009-01 Population estimates, Q1 2020. Available at: https://www150.statcan.gc.ca/t1/tbl1/en/tv.action?pid=1710000901 Accessed 30 June 2020.

6. United States Census Bureau. Annual Estimates of the Resident Population for the United States, Regions, States, and Puerto Rico: April 1, 2010 to July 1, 2019 (NST-EST2019-01), 2019. Available at: https://www.census.gov/data/tables/time-series/demo/popest/2010s-state-total.html. Accessed 30 June 2020.

7. Public Health Agency of Canada. Coronavirus disease (COVID-19): Outbreak update. Available at: https://www.canada.ca/en/public-health/services/diseases/2019-novel-coronavirus-infection.html. Accessed 8 July 2020.

8. Statistics Canada. Provisional death counts and excess mortality, January to April 2019 and January to April 2020. Available at: https://www150.statcan.gc.ca/n1/daily-quotidien/200619/dq200619b-eng.htm. Accessed 8 July 2020.

9. Weinberger DM, Chen J, Cohen T, et al. Estimation of Excess Deaths Associated With the COVID-19 Pandemic in the United States, March to May 2020. JAMA Internal Medicine, 2020; https://doi.org/10.1001/jamainternmed.2020.3391. PMID: 32609310.

10. Li R, Pei S, Chen B, et al. Substantial undocumented infection facilitates the rapid dissemination of novel coronavirus (SARS-CoV-2). Science, 2020;368:489–93.

11. Oran DP, Topol EJ. Prevalence of Asymptomatic SARS-CoV-2 Infection: A Narrative Review. Ann Intern Med, 2020; https://doi.org/10.7326/M20-3012. PMID: 32491919.

12. Skowronski DM, Hottes TS, McElhaney JE, et al. Immuno-epidemiologic correlates of pandemic H1N1 surveillance observations: higher antibody and lower cell-mediated immune responses with advanced age. J Infect Dis, 2011;203:158–67.

13. Skowronski DM, Hottes TS, Janjua NZ, et al. Prevalence of seroprotection against the pandemic (H1N1) virus after the 2009 pandemic. CMAJ, 2010;182:1851–6.

14. Skowronski DM, Chambers C, Sabaiduc S, et al. Pre- and post-pandemic estimates of 2009 pandemic influenza A(H1N1) seroprotection to inform surveillance-based incidence, by age, during the 2013-2014 epidemic in Canada. J Infect Dis, 2015;211:109–14.

15. Van Kerkhove MD, Hirve S, Koukounari A, et al. Estimating age-specific cumulative incidence for the 2009 influenza pandemic: a meta-analysis of A(H1N1)pdm09 serological studies from 19 countries. Influenza Other Respir Viruses 2013;7:872–86.

16. Skowronski DM, Moser FS, Janjua NZ, et al. H3N2v and other influenza epidemic risk based on age-specific estimates of sero-protection and contact network interactions. PLoS One, 2013;8:e54015.

17. Skowronski DM, Janjua NZ, De Serres G, et al. Cross-reactive and vaccine-induced antibody to an emerging swine-origin variant of influenza A virus subtype H3N2 (H3N2v). J Infect Dis, 2012;206:1852–61.

18. Government of British Columbia. Population projections, 2019. Available at: https://www2.gov.bc.ca/gov/content/data/statistics/people-population-community/population/population-projections. Accessed 24 June 2020.

19. World Health Organization (WHO). Coronavirus disease (COVID-19) technical guidance. The Unity Studies: Early Investigations Protocols. Population-based age-stratified seroepidemiological investigation protocol for COVID-19 infection. Available at: https://www.who.int/emergencies/diseases/novel-coronavirus-2019/technical-guidance/early-investigations Accessed 10 July 2020.

20. Health Canada. Authorized medical devices for uses related to COVID-19: List of authorized testing devices. Available at: https://www.canada.ca/en/health-canada/services/drugs-health-products/covid19-industry/medical-devices/authorized/list.html. Accessed 8 July 2020.

21. United States Food and Drug Administration. EUA Authorized Serology Test Performance. Available at: https://www.fda.gov/medical-devices/emergency-situations-medical-devices/eua-authorized-serology-test-performance. Accessed 8 July 2020.

22. Public Health England. COVID-19: laboratory evaluations of serological assays. Available at: https://www.gov.uk/government/publications/covid-19-laboratory-evaluations-of-serological-assays. Accessed 8 July 2020.

23. Theel ES, Harring J, Hilgart H, et al. Performance Characteristics of Four High-Throughput Immunoassays for Detection of IgG Antibodies against SARS-CoV-2. J Clin Microbiol, 2020; https://doi.org/10.1128/JCM.01243-20. PMID: 32513859.

24. Bryan A, Pepper G, Wener MH, et al. Performance Characteristics of the Abbott Architect SARS-CoV-2 IgG Assay and Seroprevalence in Boise, Idaho. J Clin Microbiol, 2020; https://doi.org/10.1128/JCM.00941-20. PMID: 32381641.

25. Zakhartchouk AN, Liu Q, Petric M, et al. Augmentation of immune responses to SARS coronavirus by a combination of DNA and whole killed virus vaccines. Vaccine, 2005;23:4385–91.

26. Tiwari RC, Clegg LX, Zou Z. Efficient interval estimation for age-adjusted cancer rates. Stat Methods Med Res, 2006;15:547–69.

27. Huang AT, Garcia-Carreras B, Hitchings MDT, et al. A systematic review of antibody mediated immunity to coronaviruses: antibody kinetics, correlates of protection, and association of antibody responses with severity of disease. medRxiv 2020.04.14.20065771 [Preprint]. April 17, 2020 [cited 2020 July 8]. Available from: https://doi.org/10.1101/2020.04.14.20065771.

28. Okba NMA, Muller MA, Li W, et al. Severe Acute Respiratory Syndrome Coronavirus 2-Specific Antibody Responses in Coronavirus Disease Patients. Emerg Infect Dis, 2020;26:1478–88.

29. Long QX, Tang XJ, Shi QL, et al. Clinical and immunological assessment of asymptomatic SARS-CoV-2 infections. Nat Med, 2020; https://www.nature.com/articles/s41591-020-0965-6 PMID: 32555424.

30. Payne DC, Smith-Jeffcoat SE, Nowak G, et al. SARS-CoV-2 Infections and Serologic Responses from a Sample of U.S. Navy Service Members — USS Theodore Roosevelt, April 2020. MMWR Morb Mortal Wkly Rep, 2020; 69:714–721.

31. Choe PK, CK; Suh, HJ; Jung, J; Kang, EK; Lee, SY; Song, KH; Kim, HB; Kim, NJ; Park, WB; Kim, ES; Oh, MD. Antibody responses to SARS-CoV-2 at 8 weeks postinfection in asymptomatic patients. Emerg Infect Dis [Preprint]. June 24, 2020 [cited 2020 July 8]. Available from: https://doi.org/10.3201/eid2610.202211.

32. To KK-W, Cheng VC-C, Cai J-P, et al. Seroprevalence of SARS-CoV-2 in Hong Kong and in residents evacuated from Hubei province, China: a multicohort study. The Lancet Microbe, 2020;1:e111–e8.

33. Centers for Disease Control and Prevention. Interim Guidelines for COVID-19 Antibody Testing. Available at: https://www.cdc.gov/coronavirus/2019-ncov/lab/resources/antibody-tests-guidelines.html. Accessed 8 July 2020.

34. Herzog S, De Bie J, Abrams S, et al. Seroprevalence of IgG antibodies against SARS coronavirus 2 in Belgium: a prospective cross-sectional study of residual samples. medRxiv 2020.06.08.20125179 [Preprint]. June 9, 2020 [cited 2020 July 8]. Available from: https://doi.org/10.1101/2020.06.08.20125179.

35. Pollán M, Pérez-Gómez B, Pastor-Barriuso R, et al. Prevalence of SARS-CoV-2 in Spain (ENE-COVID): a nationwide, population-based seroepidemiological study. The Lancet, 2020; https://doi.org/10.1016/S0140-6736(20)31483-5.

36. Stringhini S, Wisniak A, Piumatti G, et al. Seroprevalence of anti-SARS-CoV-2 IgG antibodies in Geneva, Switzerland (SEROCoV-POP): a population-based study. Lancet, 2020; https://doi.org/10.1016/S0140-6736(20)31304-0. PMID: 32534626

37. Doi A, Iwata K, Kuroda H, et al. Estimation of seroprevalence of novel coronavirus disease (COVID-19) using preserved serum at an outpatient setting in Kobe, Japan: A cross-sectional study. medRxiv 2020.04.26.20079822 [Preprint]. May 5, 2020 [cited 2020 July 8]. Available from: https://doi.org/10.1101/2020.04.26.20079822.

38. Xu X, Sun J, Nie S, et al. Seroprevalence of immunoglobulin M and G antibodies against SARS-CoV-2 in China. Net Med, 2020; https://doi.org/10.1038/s41591-020-0949-6. PMID: 32504052

39. Havers FP, Reed C, Lim TW, et al. Seroprevalence of Antibodies to SARS-CoV-2 in Six Sites in the United States, March 23-May 3, 2020. medRxiv 2020.06.25.20140384 [Preprint]. June 26, 2020 [cited 2020 July 8]. Available from: https://doi.org/10.1101/2020.06.25.20140384.

40. Rosenberg ES, Tesoriero JM, Rosenthal EM, et al. Cumulative incidence and diagnosis of SARS-CoV-2 infection in New York. Ann Epidemiol, 2020; https://doi.org/10.1016/j.annepidem.2020.06.004

